# Mechanistic Reorganization of Step Work in Hemiparetic Walking: Modeling and Center-of-Mass Power/Work Analysis

**DOI:** 10.64898/2026.03.11.26348174

**Authors:** Seyed-Saleh Hosseini-Yazdi, Karson Fitzsimons, John EA Bertram

## Abstract

**Purpose:** Post-stroke hemiparetic gait is commonly characterized by reduced paretic propulsion and interlimb asymmetry, yet the mechanical consequences of severely insufficient push-off for gait regulation remain unclear.

**Methods:** We examine how unilateral push-off deficiency reorganizes mechanical work across the gait cycle and regulates step length. We explored the mechanisms underlying how and why hemiparetic gait is expressed using a simple powered model and compared the model predictions against empirical data.

**Results:** The model predicts a threshold behavior: when paretic push-off falls below ~25% of optimal, compensation shifts from post-transition restoration to pre-transition energy induction, and reduced paretic step length becomes mechanically favorable, which is also seen empirically. When paretic late-stance push-off is absent, early-stance positive work emerges, vertical COM motion collapses to a single pendular motion per stride, and paretic limb step length remains disproportionately short relative to speed. As limited paretic push-off emerges, COM undergoes double pendular motion per stride, and the four-phase COM power structure previously reported in able-bodied walking reappears. Reduced single support duration and paretic positive work capacity constrains step length, which increases step length asymmetry and stance-phase energy dissipation. Paretic net COM work remains negative across speeds, whereas the nonparetic limb exhibits persistent positive net work.

**Conclusion:** Hemiparetic walking can be considered a constrained optimization problem in which decreased step length and phase-redistributed work may reflect mechanical adaptations to impaired transition mechanics, extending classical step-to-step transition and total-step work theories to pathological gait.

## INTRODUCTION

Human walking is a fundamental activity of daily living that depends on coordinated neural control, properly timed musculoskeletal force production, and whole-body mechanics. Hemiparetic gait disrupts these features^1^, yet despite extensive clinical and biomechanical characterization, a clear mechanistic explanation of ***why*** specific compensatory patterns emerge remains incomplete. Understanding these mechanisms is essential for distinguishing adaptive from maladaptive movement strategies and for guiding rehabilitation toward restoration of paretic-limb function rather than the artificial imposition of symmetry.

Walking consists of alternating single- and double-support phases. While double support represents the step-to-step transition from one limb to the next, during single support, the body’s center of mass (COM) vaults over the stance limb in a manner approximated closely by an inverted pendulum ^2–4^. Efficient walking relies on the precise timing and sequence of mechanical work performed by each limb ^5–7^. During double support, a pre-emptive push-off impulse by the trailing limb, generating the step-transition positive work, precedes the heel-strike diversion of the COM velocity by the leading limb (in dynamics a discontinuity in the COM velocity is defined as a collision, and it inevitably results in energy loss). This positive and negative work sequence has been shown to minimize the positive work required to redirect the COM during the step transition ^7,8^. During single support, early-stance rebound and late-stance preload regulate COM energy and prepare the system for the next transition ^5,6,9^. Disruption of this sequence has measurable energetic consequences ^10^: experimental reductions in ankle push-off work substantially increase contralateral collision losses and metabolic cost even at constant speed ^8,11,12^. Thus, step-to-step transition mechanics are central to walking efficiency.

### Hemiparetic Walking After Stroke, Clinical Impairment and Mechanical Consequences

Following stroke, unilateral impairments in strength, timing, and coordination lead to hemiparetic gait. Clinically, this is characterized by reduced paretic weight-bearing, prolonged and asymmetric double support, shortened paretic single support, and reduced walking speed ^13,14^. Quantitatively, paretic anterior–posterior impulse and ankle positive work are markedly reduced, with compensatory increases in non-paretic hip work ^1,15,16^.

However, the literature has largely focused on *reduced* paretic push-off work and asymmetry ^1^, without explicitly considering the mechanical consequences when paretic push-off work becomes smaller or functionally absent. Most analyses implicitly assume that some level of paretic push-off work remains available and examine how asymmetry scales from that condition, assuming that subacute hemiparetic gait is simply a failed attempt to emulate normal gait. The scenario in which paretic push-off work falls below a threshold necessary to properly participate in step-to-step transition mechanics—and how the locomotor system reorganizes under those circumstances—has not been systematically examined.

### Comparison Between Normal and Post-Stroke Walking: A Mechanistic Gap

In healthy walking, push-off, collision, rebound, and preload work are tightly sequenced within and between limbs, producing symmetric step-to-step transitions and efficient COM redirection ^6,17^. In hemiparetic walking, asymmetries are well documented, yet descriptions often remain phenomenological (identifying what changes) rather than mechanistic (why those changes are mechanically necessary or important). Predictive simulations suggest that asymmetric gait patterns under unilateral weakness can represent mechanically feasible solutions ^18^, sometimes near-optimal given the constraints of the condition. This suggests that hemiparetic gait should be understood as a separate constrained optimization problem^19^ that arrives at an alternate optimization shaped by impaired weight support and transition mechanics rather than a simple failure to achieve symmetry and emulate normal gait dynamics.

A critical but underexplored question is what occurs when paretic push-off work is insufficient to contribute meaningfully to the step transition. Step-to-step transition dynamics predicts that insufficient push-off work increases collision loss at heel-strike and necessitates compensatory positive work elsewhere in the gait sequence^OBJ^. If paretic push-off work falls below a functional threshold, the system may be forced to reorganize propulsion and support outside the normal transition window, fundamentally altering the timing and distribution of mechanical work across limbs. This regime is not explicitly addressed in current hemiparetic gait literature.

Therefore, a mechanistic framework understanding the link between impaired push-off work magnitude, weight support, and COM dynamics to the emergence of potential compensatory strategies is needed. In this study, we use GRF-derived COM work ^8^ within the framework of an extended simple powered walking model ^7,20^ considering the potential of single support power and work modulation ^6,9^ to identify how hemiparetic gait reorganizes when paretic push-off work is (i) absent, (ii) emerging, or (iii) present but limited. Rather than classifying subjects by clinical phase (sub-acute vs chronic), we propose *biomechanical phenotypes* based on the functional role of paretic push-off capacity in step-to-step transition mechanics. This approach reveals transitions in propulsion sharing and provides a mechanistic basis for distinguishing mechanical adaptations from recovery-based modifications.

## METHODS

Most of the mechanical work of walking occurs during the step-to-step transition^5^, when support alternates between limbs and the center of mass (COM) velocity must be appropriately redirected from one stance leg to the next ^5,17^. A simple model of walking suggests that the step-to-step transition is performed by the application of impulses along limbs 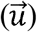 that perform mechanical work on the COM 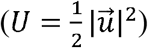^7,17,21^. This transition begins with a pre-emptive push-off impulse (*po*) by the trailing limb that partially redirects the COM velocity prior to heel strike ^17,22^. This action can be interpreted as a *generative collision* ^23^, as it alters the direction of the COM velocity vector while adding muscle produced energy to the system. The subsequent heel-strike collision impulse (*co*) of the leading limb is *absorptive* and completes the redirection of the COM trajectory to the new stance limb. A simple step-to-step transition model predicts that when push-off impulse magnitude and timing are optimal, the total mechanical work required for redirection is minimized (*W*_*post*−*transition*_~0) ^6,7^. We consider such a push-off impulse as optimal (*po*_*opt*_).

When push-off is insufficient, poorly timed, or absent, the redirection of the COM cannot be completed within the transition itself. Under such conditions, greater energy dissipation occurs at heel strike, increasing the mechanical cost of the step transition ^6,11^. We define such a push-off as sub-optimal (*po*_*sub*_). The resulting mechanical energy deficit must then be compensated for after the transition, during the subsequent single-support phase, where additional positive work is required to restore the COM energy required for continued progression ^6,9,18^. This shift of work from the transition phase into single support represents a fundamental reorganization of gait mechanics.

Based on this premise, we model hemiparetic walking using the simple walking model ^7^ under the condition of limited unilateral push-off work capacity. Specifically, we predict how the locomotor system should reorganize to optimize when paretic push-off work falls below the level required to participate effectively in step-to-step transition mechanics, and how compensation is redistributed across phases of the gait cycle. We then compare these modeled predictions to empirical gait measurements in stroke survivors.

### Simulations and Predictions

Since it has been shown that most of the step work occurs during the step-to-step transition ^7^, first we focus on the step-transition assuming passive COM motion during single stance ^3,4,7^. When post step-transition mechanical energy compensation is required, we assume the work duration is very short ^6,7^. It has been shown that hemi-paretic walking coincides with reduced propulsion performance on one side stemming from deficiencies in ankle joint actuation ^1^. As such, we utilize a simple walking model ^7^ to evaluate the consequence of reduced push-off work

^18^. The model has massless rigid legs and concentrated mass at the pelvis ^7^. We adopted this model since it can successfully predict the optimal strategies for walking over complex terrain ^24^, predict the preferred step length for a given walking speed ^20^, and predict the preferred joint to compensate for step-transition mechanical energy deficits ^9^ despite its simplicity and lack of anatomical detail. It can also indicate mechanically preferred walking speeds ^6^, and evaluate the mechanical cost of shortened step lengths. **TABLE *1*** summarizes the simulation parameters and assigned magnitudes and the way they are calculated.

**TABLE 1.** Simulation parameters and variables used in the simple walker model. The magnitude column reports either the assigned simulation value or the analytical relationship used to determine each variable.

| Parameter | Description | Magnitude |
| --- | --- | --- |
| $po$ | Push-off impulse | $po_{opt} = v_{ave} \cdot \tan \alpha$ |
| $PO$ | Push-off work | $\frac{1}{2} po^2$ |
| $po_{max}$ | Maximum push-off impulse | $v_{ave} \tan 2\alpha$ |
| $co$ | Collision impulse | $co_{opt} = po_{opt}$ |
| $CO$ | Collision work | $\frac{1}{2} co^2$ |
| $v_{ave}$ | Average walking speed | 1.0 m/s |
| $v^-$ | Pre-step-transition velocity | $v^- \sim v_{ave}$ |
| $v^0$ | Mid-step-transition velocity | $v_{opt}^0 = \frac{v_{ave}}{\cos \alpha}$ |
| $v^+$ | Post-step-transition velocity | $v_{opt}^+ = v_{ave}$ |
| $l$ | Leg length | 1.0 m |
| $m$ | Mass | 1.0 kg |
| $2\alpha$ | Inter-limb angle between leading and trailing leg at step-transition point | $\alpha = \frac{S}{2}$ |
| $\gamma$ | COM partial redirection due to sub-optimal push-off | $\gamma = \arctan\left(\frac{po_{sub}}{v_{ave}}\right)$ |

For the sake of simulation, we present the impulses and work normalized by mass (*m* = 1 *kg*). Thus, the impulse and work units become *m*.*s*^−1^ and *J*.*kg*^−1^, respectively^7,21,25^. With small angle approximation, we assume that unperturbed pre-step-transition COM speed is equal to average step speed: *v*^−^~_*ave*_. We also approximate the step length by the inter-limb angle (2 *α*) while all step-transition impulses are along the associated limbs^7^.

At the step-to-step transition, if the push-off work for a certain walking speed (*v*_*ave*_) is optimal (where the entire positive step work occurs at the step transition, 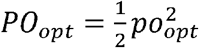), the magnitude of the collision loss will be equal to the push-off work ^7^. As such, the post-transition speed will not change (**FIGURE 1**A). However, when the push-off impulse is suboptimal (*po*^*sub*^ < *po*_*opt*_), it cannot completely adjust the COM velocity trajectory before the subsequent heel-strike (**FIGURE 1**B) ^17^. As a result, the direction change of the partially modified COM velocity is ^6^:

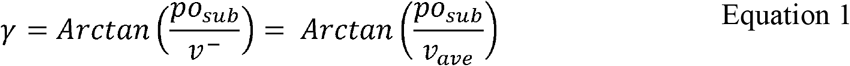

**FIGURE 1.**
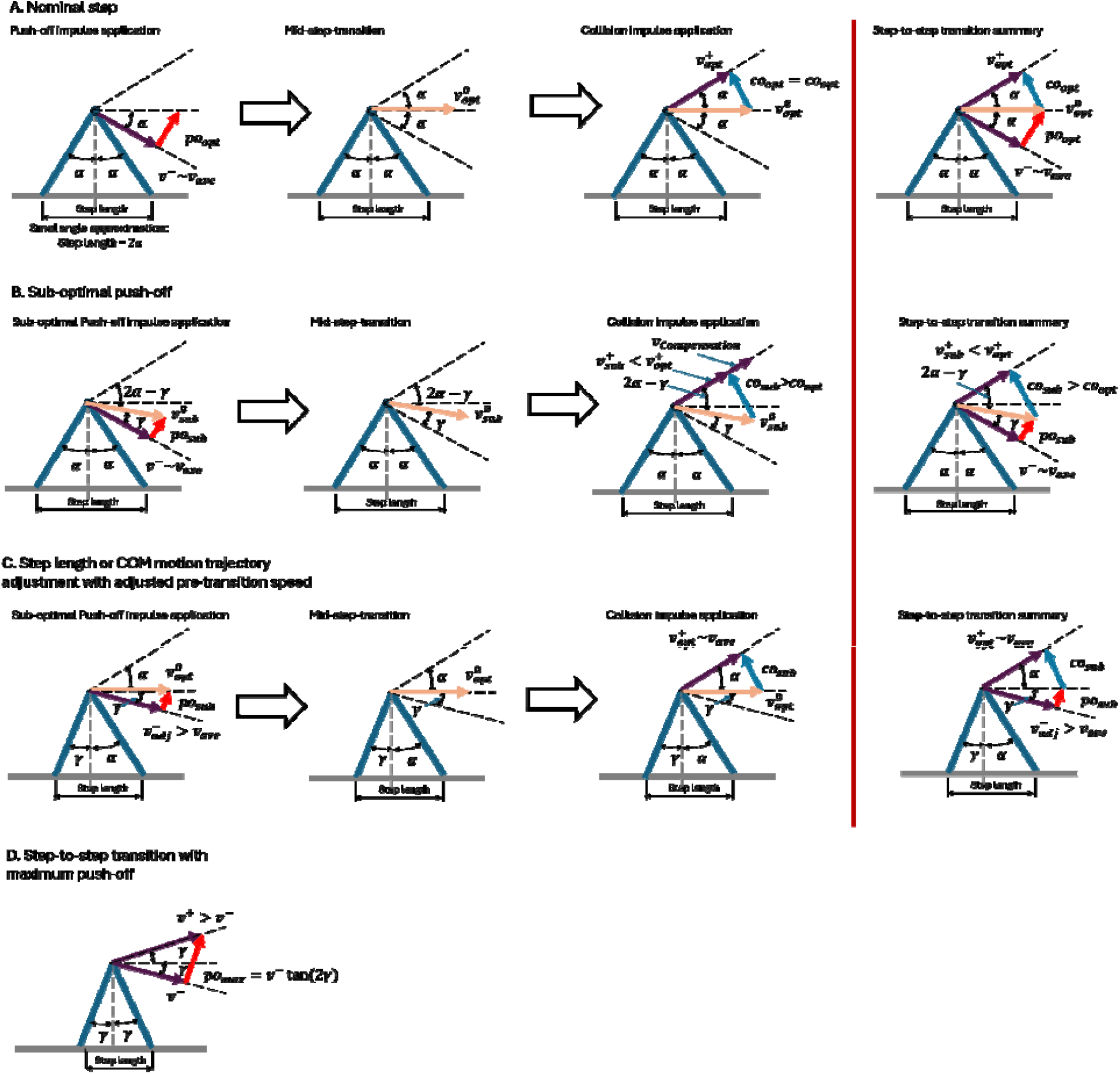
Analytical simulation of walking based on a simple powered model: (a) optimal step-transition for a prescribed walking speed for which the optimal push-off and collisions are equal, (b) when push-off is less than optimal (, the subsequent collision dissipations grows and as such the post transition speed declines, (c) step length o COM motion trajectory maybe adjusted to match the magnitude of sub-optimal push-off to achieve prescribed average walking speed post-step-transition (d) The step transition may be performed when the trailing leg provides the entire transition work demand with maximum push-off.

If 2*α* represents the trailing and leading legs’ angle at the point of transition, as 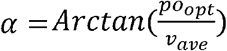, and since *po*^*sub*^ < *po*_*opt*_, thus, *γ* ^*sub*^ < *γ*_*opt*_. We show the mid-transition COM velocity by 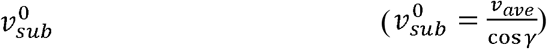, therefore, the subsequent heel-strike collision impulse required to align the COM trajectory to the next step and the post transition speed 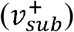 will be calculated as ^6^:

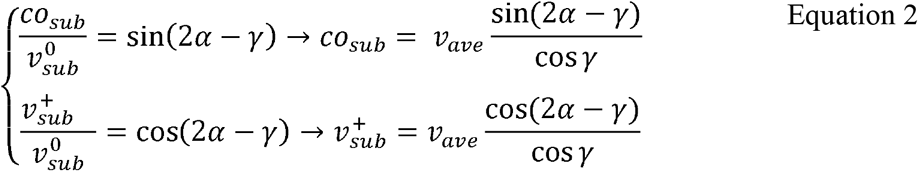

Since *γ* > *γ* → 2*α* > 2*γ* − *γ* > *γ*, therefore, cos (2*α* − *γ*) < cos *γ*, and as such, 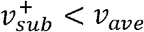. It means that when a walker exerts a suboptimal push-off impulse, the associated heel-strike dissipation increases and hence its post-transition speed declines (FIGURE 1B). To maintain a constant average walking speed (e.g. treadmill walking) post-transition, extra positive energy must be added.

Since older adults are more prone to strokes and degenerative conditions, we run the simulation for *v*_*ave*_ = 1.0 m.s^−1^ for which *α* = 0.3 rad. This is suggested as the mechanically preferred speed for older adults ^6^.

For example, when the walker exerts only 80% of the optimal push-off impulse, the post step-transition speed becomes 0.77 m.s^− 1^, which requires 0.02 J.kg^− 1^ energy makeup in the form of compensation. Instead of post step-transition mechanical energy compensation, the walker might also choose to compensate for a suboptimal push-off impulse by adding extra positive mechanical energy before the step transition, e.g. through hip actuation ^8,18^. Since subsequent to the deficient push-off impulse, heel-strike occurs in the unaffected leg, we assume that its impulse and energy loss are optimal (*co*_*opt*_ and *po*_*opt*_, **FIGURE 1**C). Hence, the issue is to find the pre-transition compensation required to sustain momentum post step transition 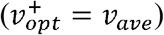. The optimal collision impulse is equal to the optimal push-off (*co* _*opt*_ = *po* _*opt*_), thus:

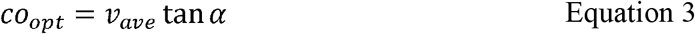

Therefore, the angle between *v*^0^ and *v*^+^ must be equal to *α*. This means that the mid-transition speed approaches optimal walking. Accordingly, we calculate the magnitude of the mid-transition COM velocity knowing the post transition speed 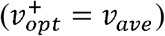 as:

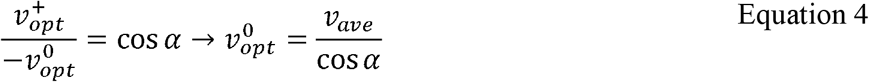

On the other hand, when the push-off impulse is deficient (sub-optimal), the redirection angle of the COM velocity vector is reduced (*γ* < *α*, Equation 1). Therefore, the pre-transition COM velocity magnitude required to result in the prescribed walking speed post step-transition must be:

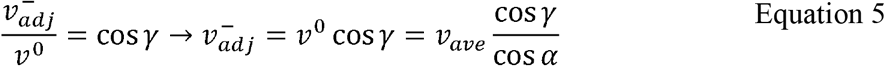

This means that the trajectory of the COM motion must be modified in a way that 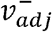 makes angle of *γ* with the horizontal while its magnitude is also adjusted by pre-transition positive energy addition. **FIGURE *2*** compares the post and pre-transition mechanical energy and the total positive mechanical work required when a suboptimal push-off impulse as a fraction of the optimal push-off is applied. As demonstrated (**FIGURE 2**C), there is a threshold for which the total mechanical work of pre-transition compensation exceeds the post-transition compensation strategy (~0.25 *po*_*opt*_). We can use this threshold as a criterion to distinguish the severity of the deficit biomechanically. Therefore, when the pre-emptive suboptimal push-off is less than 0.25 *po*_*opt*_, a pre-transition compensation becomes mechanically favorable. Thus, we expect positive work (*W*^+^) to be implemented before the step-transition. However, if *po*_*sub*_ > 0.25 *po*_*opt*_, the positive mechanical compensation must be added after the step-to-step transition. This will be represented by an increase in the rebound work of the contralateral limb^1,26^, where rebound work is suggested to be association with hip work ^8,27^

**FIGURE 2.**
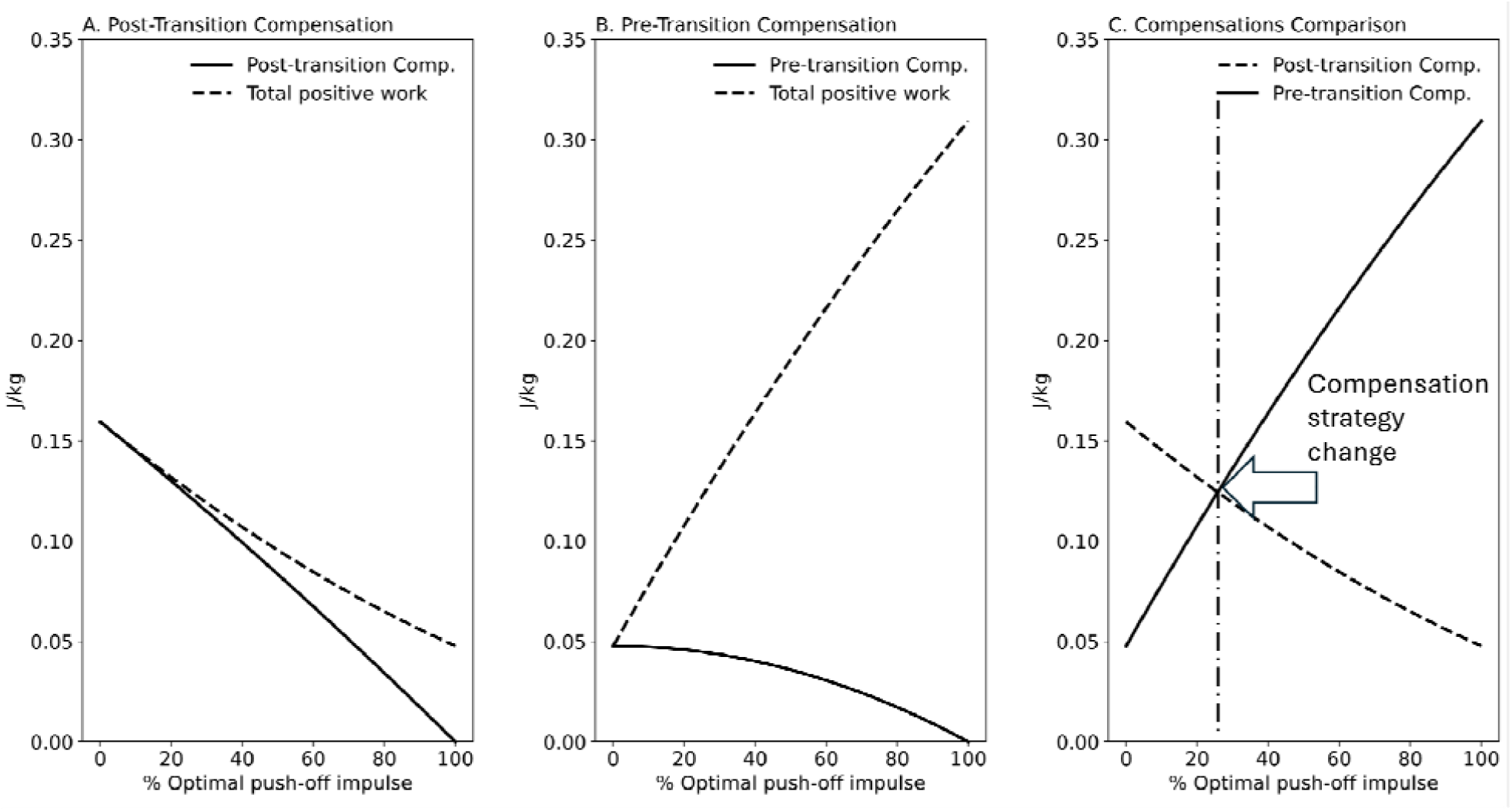
Step transition energy compensation and total mechanical work when the pre-emptive push-off ranges from zero to the optimal magnitude for a prescribed walking speed: (a) post transition strategy in which the mechanical energy deficit is compensated after the step-to-step transition, (b) pre-transition strategy in which the COM speed is increased before the step transition by adding mechanical energy to compensate for the energy dissipation of heel-strike (step-transition collision), (c) comparison of total step work performance for each strategy, i.e. post step transition energy compensation vs pre-step transition compensation: when the pre-emptive push-off is less than 25% of the optimal push-off for a given walking speed, the pre-step-transition energy infusion is preferable whereas if the pre-emptive push-off exceed 25% of the optimal magnitude, the post-step-transition strategy becomes favorable mechanically. The 25% of optimal push-off threshold might be used to categorize the walkers’ condition biomechanically.

For a suboptimal push-off impulse, the extreme case occurs when *po*_*sub*_ = 0 → *γ* = 0, and thus the pre-transition COM speed must be increased to 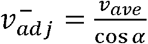 locally by adding positive work to maintain average walking speed following the step-transition.

The modification of the COM motion trajectory might be achieved by shortening the step length. It is shown that the mechanical work demand increases exponentially with step length when walking speed is kept constant^5^. Therefore, reducing step length may reduce the mechanical demand for COM trajectory redirection^17^. **FIGURE 3** shows that with an increase of step length from 0.61 m to 0.82 m, the required optimal push-off work (push-off impulse: po = *v*_*ave*_.tan *α*, and associated push-off work: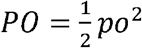) increases from 0.03 J.kg^−1^ to 0.24 J.kg^−1^. For step length of 0.7 m, the required nominal push-off is 0.08 J.kg^−1^. When the applied push-off ranges from 0 J.kg^−1^ to 0.08 J.kg^−1^, the required compensation decreases from 0.25 J.kg^−1^ to 0. As a result, the total step work declines from 0.25 J.kg^− 1^ to 0.08 J.kg^− 1^.

**FIGURE 3.**
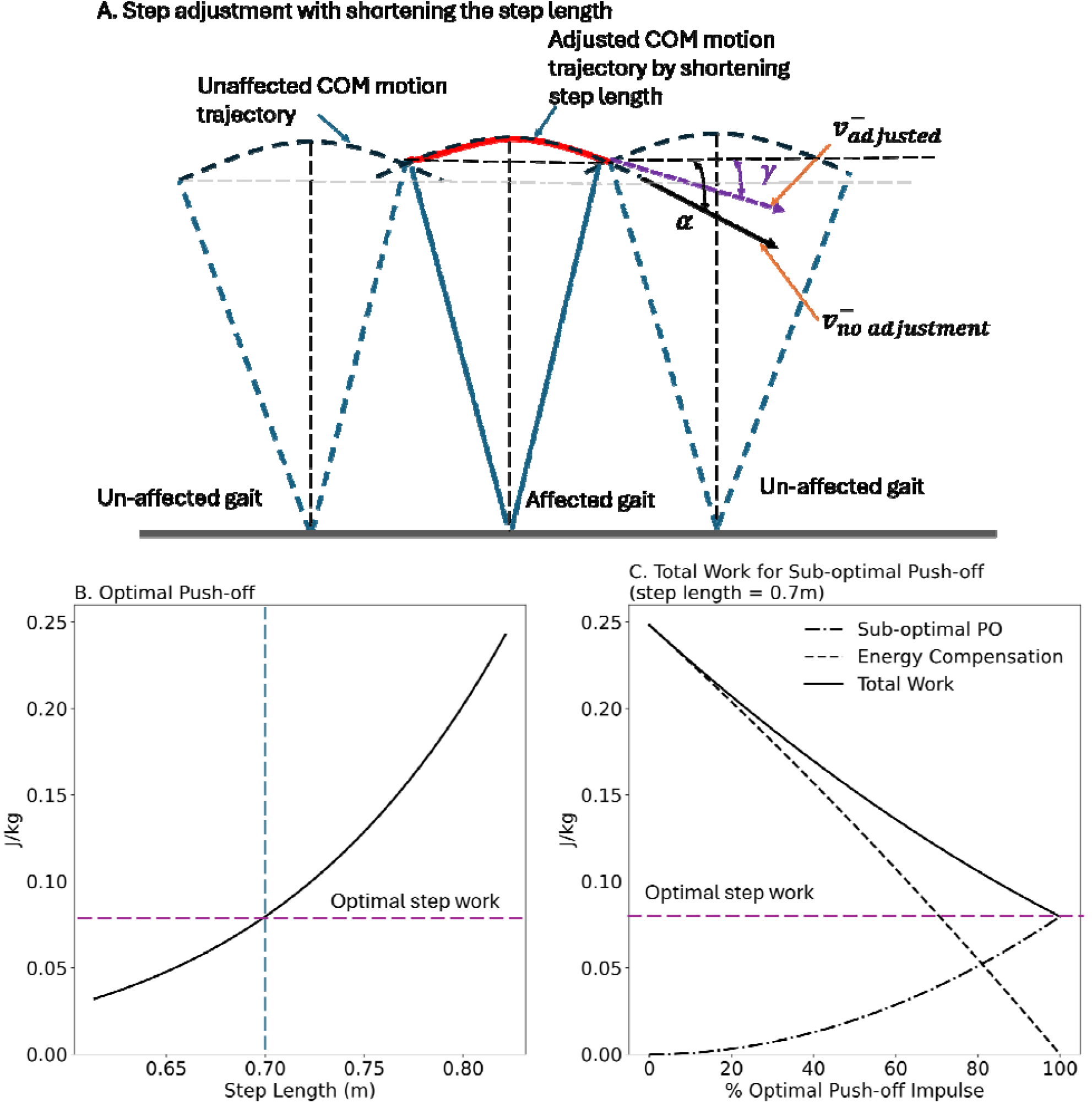
A post stroke walking may be divided into an unaffected gait with optimal push-off and affected gait with a suboptimal push-off for the imposed walking speed. The affected side is not able to produce the same amount of positive work for the step transition, (). Since the step transition work is proportional to step length, one strategy is to reduce the step length at the paretic side to match its work performance capacity. As a result, the COM motion trajectory and direction of its velocity become flatter (a). (b) the analytical optimal push-off work for given step length. (c) When the push-off work is lower than optimal magnitude, energy must be added to increase the total step work and maintain momentum.

Since it is suggested that nominal walking single support (following an optimal step-to-step transition) resembles the motion of an inverted pendulum, walking with shorter step length will truncate the arc which will approach a flatter COM path. In the extreme case the COM path will asymptote to horizontal^28^. As such, single support motion may be represented by a linear inverted pendulum model ^29^.

Another mechanism for modulating COM velocity is adjustment of the stance limb’s effective length during single support. Muller et al^30,31^ have shown that humans adjust their effective leg length when they face terrain obstacles. By altering leg extension or flexion, the system can influence the vertical and horizontal components of COM motion. When the COM trajectory approaches horizontal, the rigid leg assumption is relaxed. Nonetheless, this does not impact our simulation since we retain the rigid-leg approximation during the brief step-to-step transition (**FIGURE 4**) when the load is distributed between the two limbs. Under this reduced individual-limb loading condition, we assume that the paretic limb provides sufficient effective rigidity for the transition to be approximated using the rigid-limb model.

**FIGURE 4.**
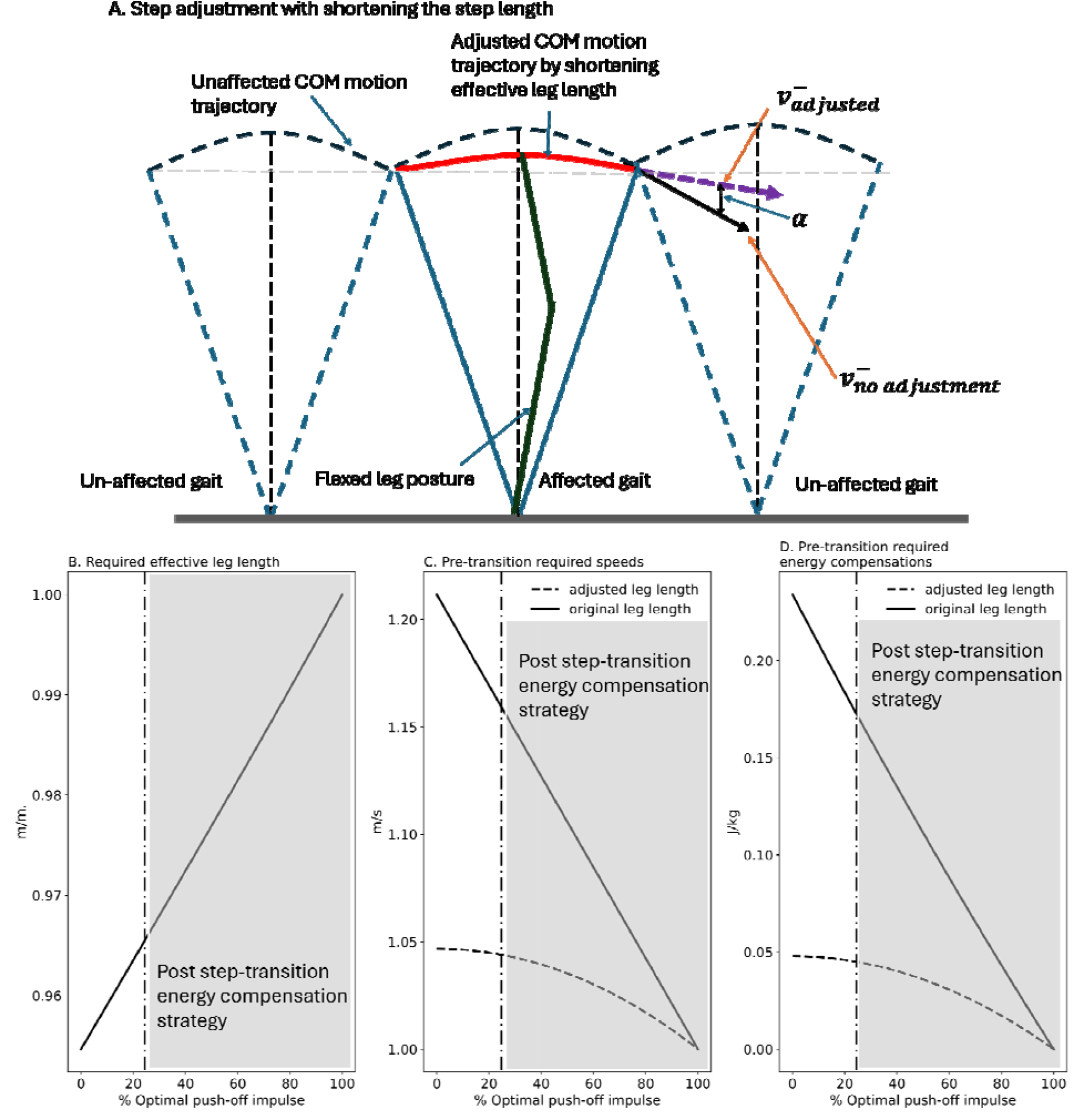
(a) Modulation of COM motion trajectory may be attained by adjusting the effective leg length. As a consequence, the step-transition positive mechanical work demand is reduced. (b) the adjusted leg length based on the available push-off capacity, (c) pre-transition adjusted walking speed for adjusted leg length to attain the prescribe walking speed post step-transition, (d) pre-transition energy compensation to modify the COM speed before step-transition.

For the sake of this simulation, we focus only on a push-off impulse magnitude of less than 25% of optimal magnitude for given walking speed. As the available push-off impulse rises from 0 to 0.25 of optimal push-off magnitude, the required effective leg length also increases from 0.95 to 0.96 of original leg length (assumption: original leg length is 1m). Assuming the average walking speed of 1.0 m.s^−1^, when the effective leg length is not adjusted, the pre-transition compensation ranges from 0.23 J.kg^−1^ to 0.17 J.kg^−1^ to achieve pre-transition speed of 1.21 to 1.16 m.s^−1^. Whereas, when the effective leg length is adjusted, the pre-step transition compensation only ranges from 0.05 to 0.04 J.kg^−1^ to reach 1.05 to 1.04 m.s^−1^ just before step transition.

We expect that the paretic limb generates substantial negative work because of its deficits. The reduction of the paretic side’s step length decreases the step-transition work demand. Hence, it is fair to expect that if the step length reduction is to match paretic positive work capacity, the paretic side’s net step work becomes close to zero 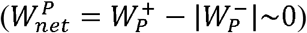. Therefore, extra paretic step length reduction may stem from mechanical energy dissipation outside of the step-transition phase, i.e. single support, leading to 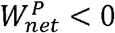.

Therefore, we also quantify the impact of negative work during single support. We use a pendular system for single support^8^ with a constant opposing hip torque. We assume the initial walking speed as 1 m.s^−1^ for step length of 0.6 m. Two conditions are evaluated: opposing torque applied from the beginning of single support and opposing torque initiated at mid-stance. For each condition, we quantify the corresponding opposing mechanical work, single-support duration, and achievable step length. We observe that, when an opposing torque is applied, the step duration increases as the COM angular velocity declines. However, this relationship is not continuous. Beyond a critical opposing-work magnitude, COM angular velocity reaches zero before completion of the prescribed step, indicating failure of forward pendular progression. We assume that such a situation warrants an advanced heel-strike to avoid falling backward or collapsing. Hence, the achievable step length and duration are reduced. The critical opposing-work magnitude depends strongly on the timing of energy removal, occurring at approximately 0.12 J when opposing work begins at single-support onset and 0.33 J when it begins at mid-stance. Thus, the model predicts a piecewise response in which increasing energy dissipation initially prolongs single-support progression, but beyond a timing-dependent energetic threshold, both step duration and achievable step length decline sharply. (**FIGURE 5**).

**FIGURE 5.**
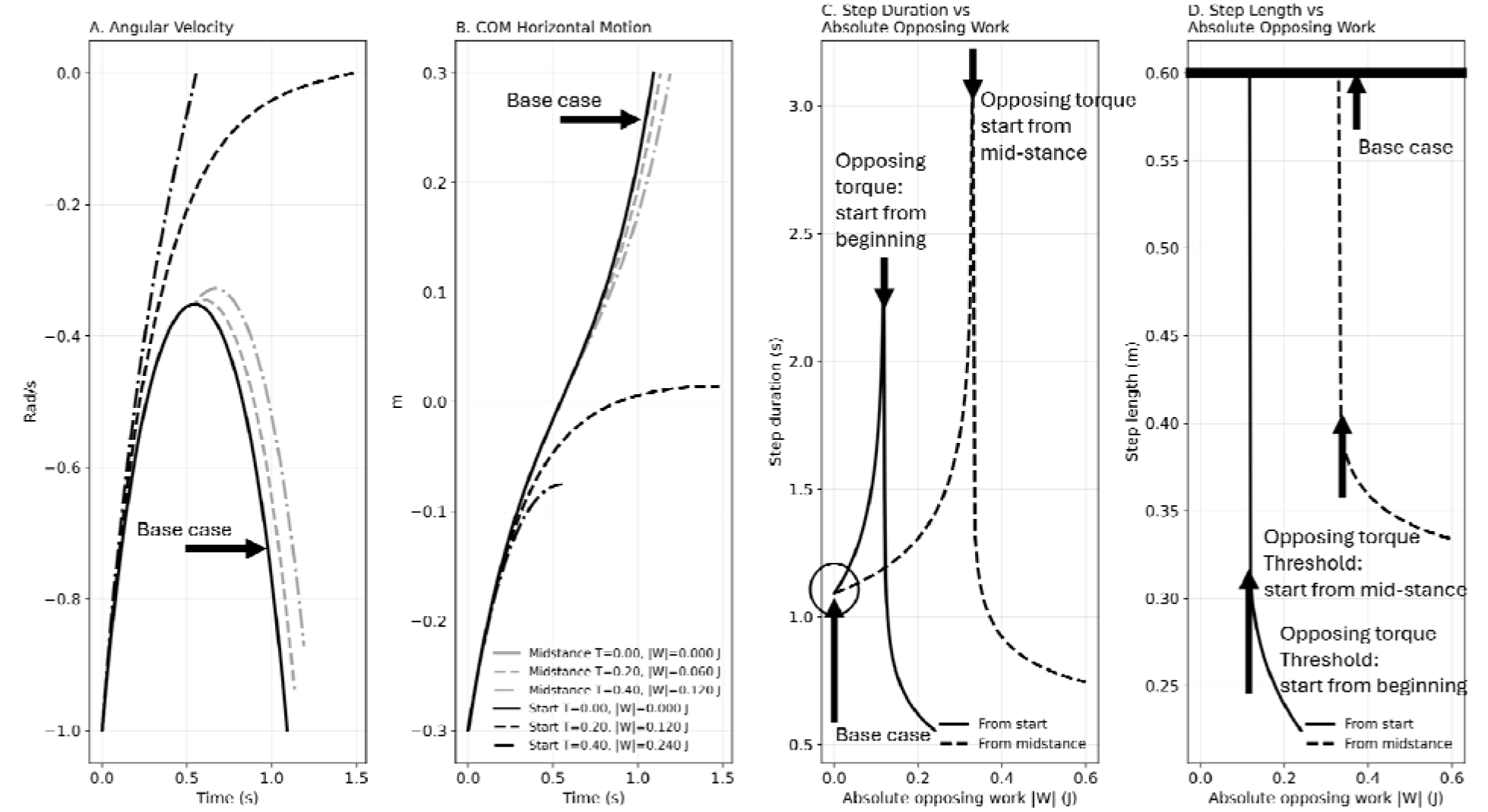
Single-support progression in an inverted-pendulum model subjected to opposing mechanical work. (a) COM angular velocity and (b) COM horizontal travel for opposing torque applied from single-support onset (black) or from mid-stance (gray). (c) Single-support duration and (d) achievable step length as functions of opposing work. Increasing opposing work initially prolongs single-support duration; beyond a timing-dependent critical work magnitude, angular velocity reaches zero before completion of the prescribed step and both duration and achievable step length collapse. The critical opposing-work magnitude is approximately 0.12 J for opposition beginning at stance onset and 0.33 J when opposition begins at mid-stance.

Based on the simulation results we propose three predictions as hypotheses to be tested by empirical data:

1. Hemiparetic gait exhibits a regime shift in the timing of positive mechanical compensation governed by paretic push-off work capacity. When paretic push-off work is severely reduced (posub ≪ po_optimal_), compensation will occur before the step-to-step transition; whereas, when paretic push-off work is moderately reduced (posub < po_optimal_), compensation will occur after the transition.
2. When paretic push-off work capacity is suboptimal, an individual will reduce paretic step length to match the trailing-limb push-off impulse capacity at the paretic-to-unaffected transition
3. To adjust the step-transition mechanical energy demand, the COM motion trajectory might also be adjusted to reduce positive work demand.
4. If the paretic step length reduction does not cover the step-transition mechanical work demand, i.e. 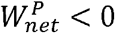, there must also be extra energy losses outside of the step transition phase. Therefore, with net negative work dominance during paretic stance, the non-paretic limb will show net positive work dominance, reflecting an energy dissipation in paretic limb and energy generation on the unaffected side.

### Empirical Data and Hypothesis Evaluation Study Participants

Eleven individuals (N = 11, 5 females and 6 males, age 54.6±11.5 (SD) years, mass 78.03±18.42 kg, height 168.79±12.39 cm, 10.0±7.0 weeks post-stroke at first assessment) undergoing post-stroke inpatient rehabilitation were recruited as part of a larger study on the impacts of split-belt treadmill walking during rehabilitation. Participants were included if they were over the age of 18 and had experienced their first ischemic or hemorrhagic stroke within the past 6 months. Participants were excluded if they had other history of significant neurological injury or disease, lower limb orthopedic issues, excessive pain that impacted walking ability, and if they had experienced a cerebellar stroke. Individuals were recruited from the Neuro Rehab Unit of the Foothills Hospital in Calgary, Alberta, CA. Potential participants were identified by relevant clinicians working on the unit. The study team approached potential participants for recruitment and obtained written informed consent according to protocol approved by the Conjoint Health Research Ethics Board at the University of Calgary (Ethics ID REB21-1576).

### Treadmill Setup

Participants walked on a split belt instrumented treadmill (Bertec Corp, Columbus Ohio) with embedded force plates recording ground reaction force (GRF) at 1000Hz. Each participant wore a harness connected to a patient lift capable of fall arrest. A semi-immersive virtual reality pathway was displayed across screens in front of the treadmill, with the visual flow of the pathway matched to the speed of the treadmill.

### Data collection protocol

Baseline comfortable walking speed was established via the mean of two bouts of the 10 m walk test with the use of gait aids (canes, walkers, etc.) permitted. Heart rate and blood pressure were measured before and after each walking bout to ensure cardiovascular safety of participants. Participants then completed at least 2 minutes of treadmill walking at a safe speed selected by their physiotherapist. During treadmill trials participants were not permitted to use walking aids but were allowed to lightly touch an anterior handrail and receive coaching from their physiotherapists. Participants completed multiple bouts of treadmill walking over the course of their inpatient rehabilitation, with up to 3 sessions per week and up to 13 sessions total.

Treadmill speed was increased over the course of the assessments at the discretion of the participants’ physiotherapists. All study protocols were completed in a way that minimized interference with the participants prescribed rehabilitation and occurred within the same facility as their clinical rehabilitation activities. To minimize the influence of body-weight support provided by the safety harness, handrail use, or therapist assistance, only trials for which the mean body weight estimated from the vertical ground reaction force differed by less than 5% from the independently measured body weight were included in the analysis. Trials exceeding this threshold were excluded. For trials meeting this criterion, individual strides were additionally screened for incidental external weight support. For each stride, the mean total vertical ground reaction force, calculated from the summed vertical forces of both limbs over the complete stride, was compared with the independently measured body weight. Individual strides differing by more than 5% from measured body weight were excluded as outliers.

### Empirical Data Analysis

To test our hypotheses, we analyze walking ground reaction forces (GRFs) to derive limb-specific COM mechanics. First, raw GRFs are low-pass filtered (3rd order, 10 Hz cutoff^32^) to remove high-frequency noise. The filtered GRFs are summed and divided by body mass to compute COM acceleration, assuming steady-state walking during extended data collection beyond a single stride ^5,26^. The COM velocity is then obtained by time integration of the acceleration. To remove integration drift, the computed COM velocity is high-pass filtered with zero phase shift (3rd order, 0.256 Hz cutoff) ^5,10,26^. We used the COM speeds to compute the COM positions. For vertical direction, the COM position was demeaned (detrended). Vertical COM excursions were identified qualitatively from the population-average COM trajectory; excursion number was not quantified at the individual-participant level.

Using the individual-limbs method ^5^, limb-specific COM power profiles are computed 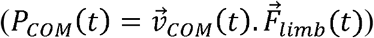 and ensemble-averaged to generate representative hemiparetic walking patterns for paretic and unaffected sides. Since the push-off is defined as the positive COM work occurring during late-stance double support ^8^, when a distinct push-off phase is absent in the COM power profile, we examine whether positive step work is redistributed to early stance, which we interpret as pre-step-transition positive work redistribution as predicted by the model. We calculated the total step’s positive (*W*^+^) and negative work (*W*^−^) by time-integrating COM power profile when *PCOM* (*t*) > 0 and *PCOM* < (*t*) 0, respectively.

Vertical GRF events are also used to detect gait phases and calculate the paretic and unaffected step lengths and durations. We then evaluated how interlimb step length varies across walking speeds. Finally, total positive and negative work are computed to derive net COM work for each specific limb, and their speed-dependent changes are analyzed. Speed-dependent relationships were evaluated using linear mixed-effects models, with participant included as a random effect to account for repeated measurements across walking speeds.

## RESULTS

Ground reaction force data were analyzed across five walking speeds (0.2–0.7 m.s^−1^) to quantify temporal parameters and limb-specific COM work. We also calculated the COM limb specific average power trajectories to investigate how the gait pattern was generated and examine differences from COM work patterns previously reported in able-bodied walking (**FIGURE 6**).

**FIGURE 6.**
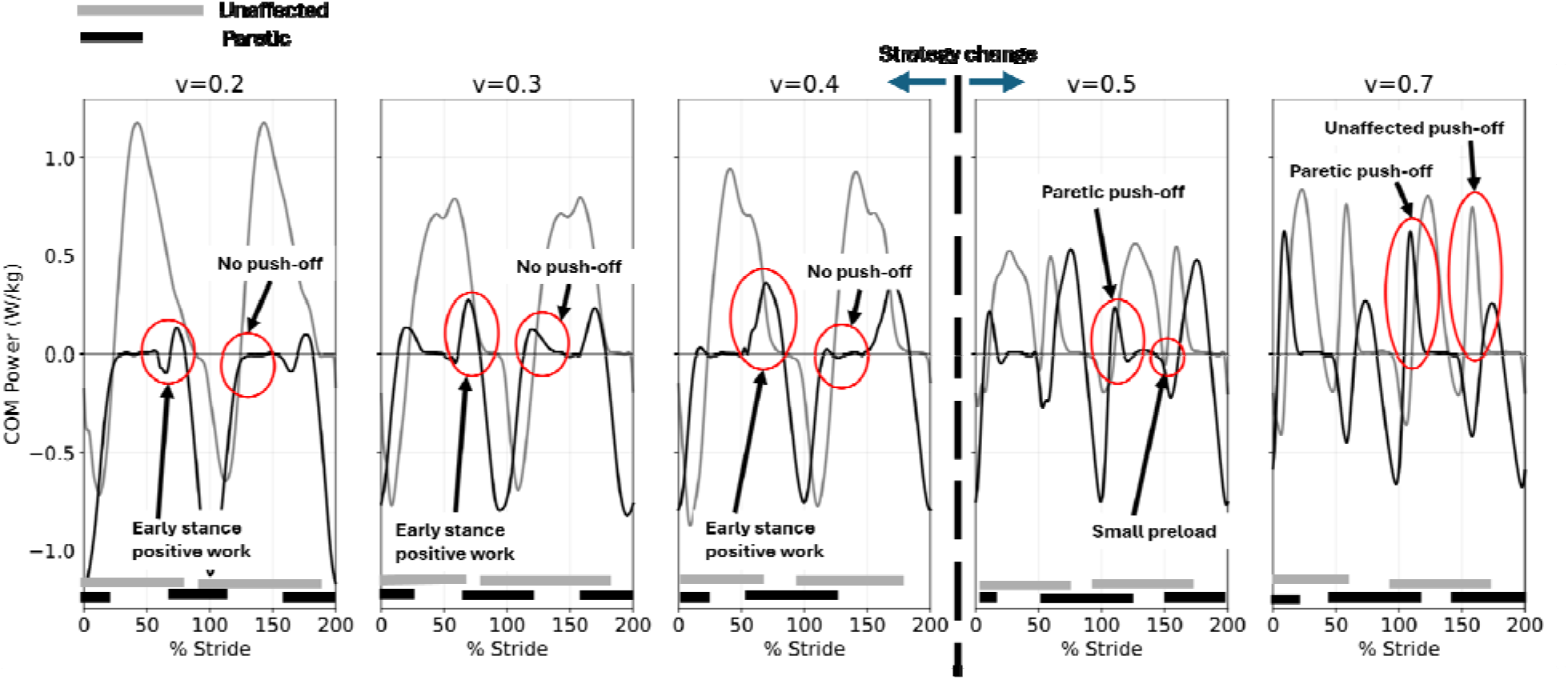
The COM power trajectories for paretic (Black) and unaffected side (gray) as the walking speed increases. At lower speeds and on the paretic side, there is no visible push-off, and the positive step work occurs at the beginning of stance, mainly during double support. As walking speed increases, positive work is also extended to the subsequent single support. With walking speed increase (0.5 ^-1^ paretic side appears that grows with speed.

At low walking speeds (*v_ave_*< 0.5 m.s^−1^) it was not possible to distinguish push-off in late stance on the paretic side, yet positive work occurred in early double support. With speed increase, the positive work was extended into the subsequent single support. We could also detect a small collision in early stance for 0.2 that decreased as walking speed increased (0.3 and 0.4). The main negative step work, however, was during single support that extended to toe-off. At 0.5 would observe a push-off that grew with speed.

On the unaffected side, the step started with a heel-strike collision that was followed by a large addition of positive work that lasted until toe-off. Therefore, we did not observe the typical four-phase COM power profile ^8^. In normal unaffected walking, COM power starts with negative power at heel-strike (collision), changing to positive power in early single support (rebound) shifting to negative power in late single support (preload), followed by a positive power push-off in late double stance. When the paretic limb was able to perform push-off, the four-phase COM power profile emerged, initially with quite small preload on the unaffected side. With speed increase, both sides showed COM power profiles that more closely resembled COM power profiles previously reported in able-bodied walking^8^.

The COM vertical motion trajectory showed a speed-dependent change in COM oscillation form. At walking speeds greater or equal to 0.5 *m.s*^− 1^, the vertical COM displacement exhibited two distinct oscillations per stride, characterized by alternating minima at each heel-strike and intermediate maxima near mid-single support, consistent with the healthy pendular pattern of human walking. In contrast, at lower walking speeds (<0.5 *m.s*^−1^), only a single rise–fall oscillation per stride was observed. Specifically, unaffected-heel strike occurred close to the vertical COM minimum, followed by a monotonic rise during unaffected stance to a maximum coincident with paretic heel-strike, after which the COM descended continuously until the subsequent unaffected heel-strike. At lower speeds, no secondary peak in vertical displacement was observed during paretic stance. Accordingly, the vertical COM trajectory exhibited one rather than two pendular excursions per stride, corresponding to the absence of a distinct pendular excursion during paretic single support at the lower walking speeds. This indicates a structural alteration in stride-level COM dynamics when the paretic push-off was absent (**FIGURE 7FIGURE 7**).

**FIGURE 7.**
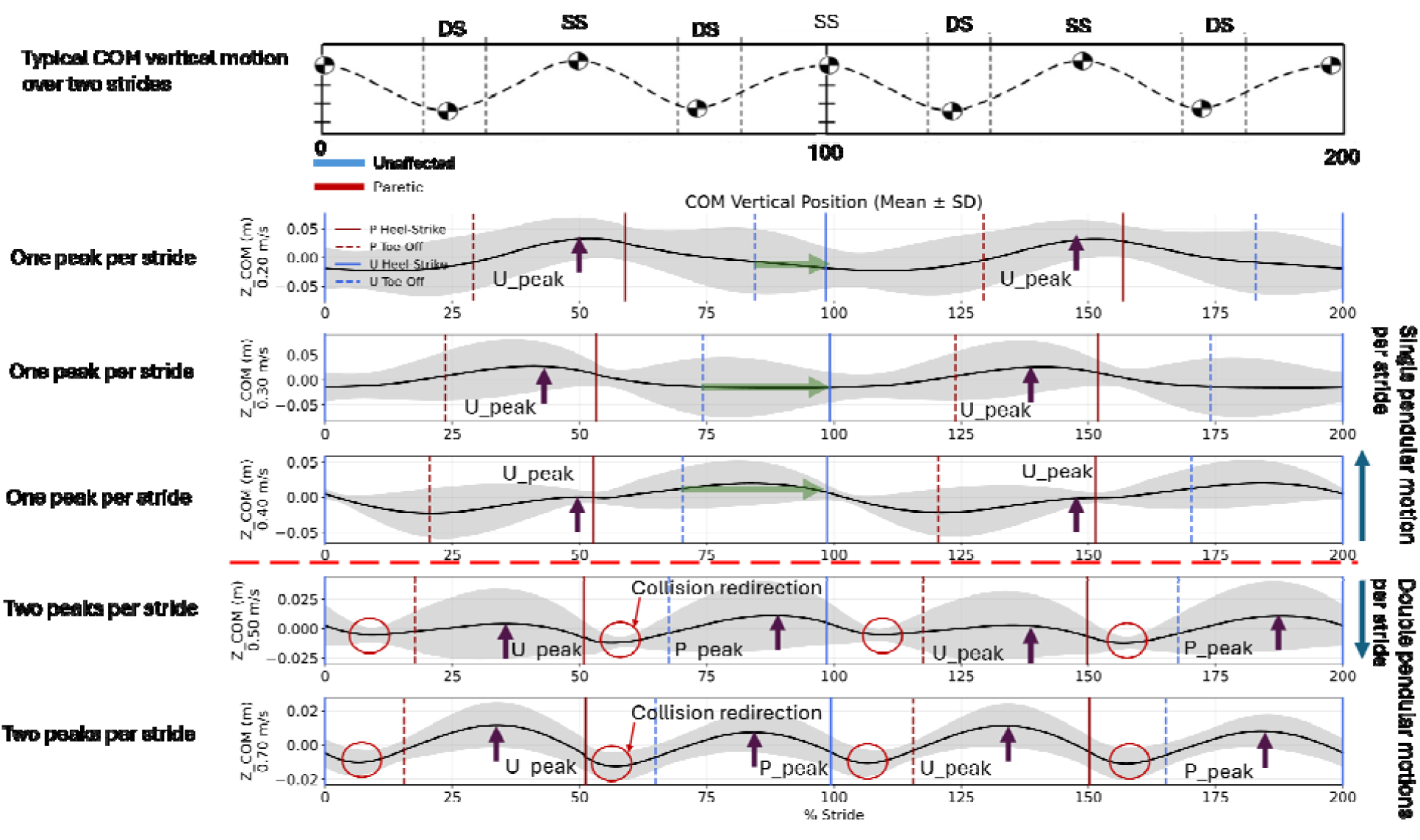
COM motion trajectory over two consecutive strides. The typical COM vertical motion indicates two pendular-like motion for each limb with their associated peaks over one stride. At lower speeds () the paretic COM motion depicted a flattened trajectory indicated by green arrows. It also did not show the heel-strike collision COM motion redirection for the paretic side. Therefore, it does not indicate redirection at unaffected side to paretic transition. With higher walking speeds () we observe that both limbs demonstrate pendular motion with their associated COM vertical peak during single support, and COM redirections are evident in double support phases.

All speed-dependent associations and interlimb relationships were evaluated using **linear mixed-effects models (LMMs)**, with participant included as a random effect to account for repeated measurements. The outcomes included total step positive COM work (), total step negative COM work (), step net COM work (), single-support net COM work (), single-support duration, and step length (**FIGURE 8** and TABLE ***2***).

**FIGURE 8.**
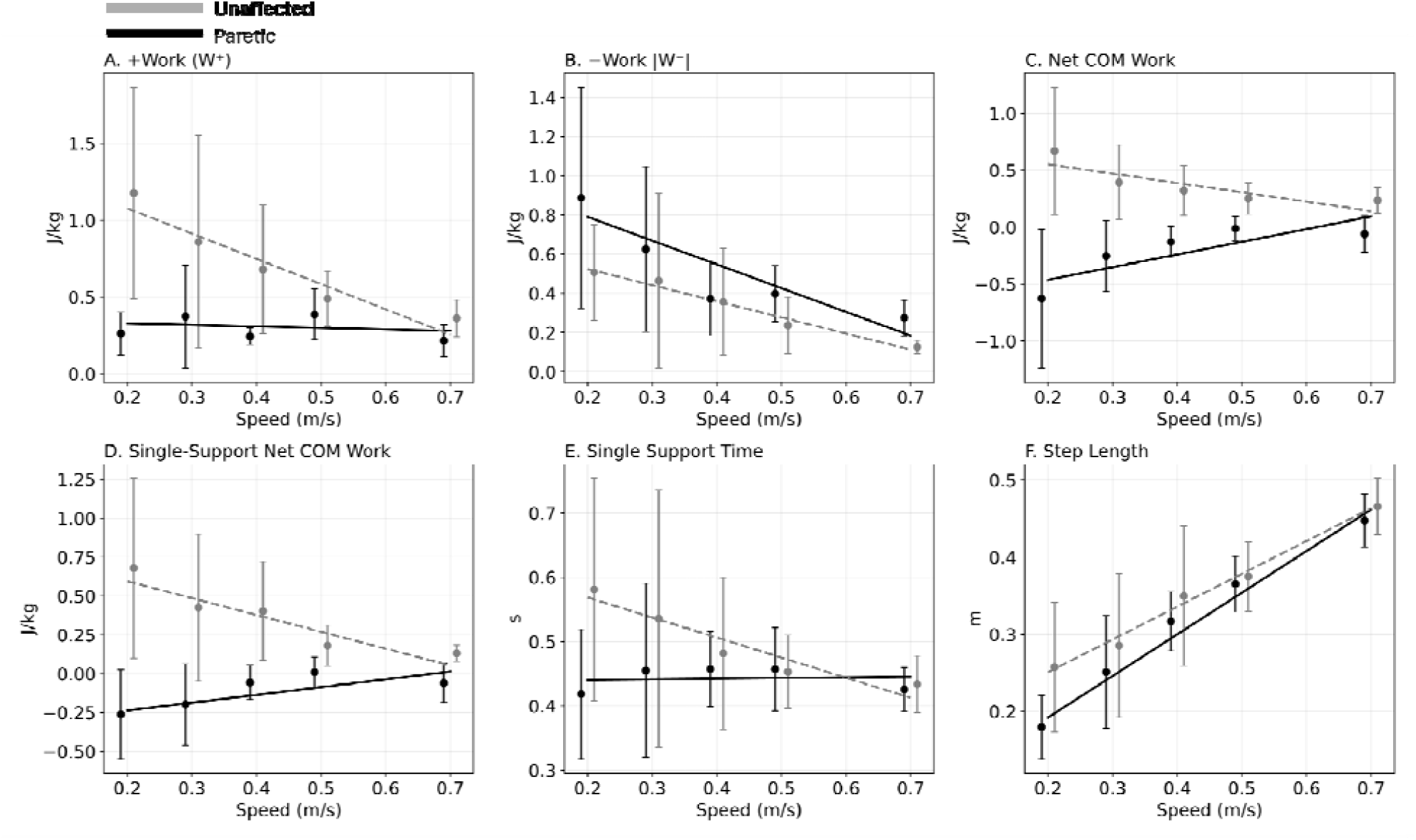
Hemi-paretic walking gait parameters and metrics for unaffected (grey) and paretic (black) sides: (a) total step positive work, (b) total step negative work, (c) total net step work, (d) net single support work, (e) single support duration, and (f) step length.

**TABLE 2.**
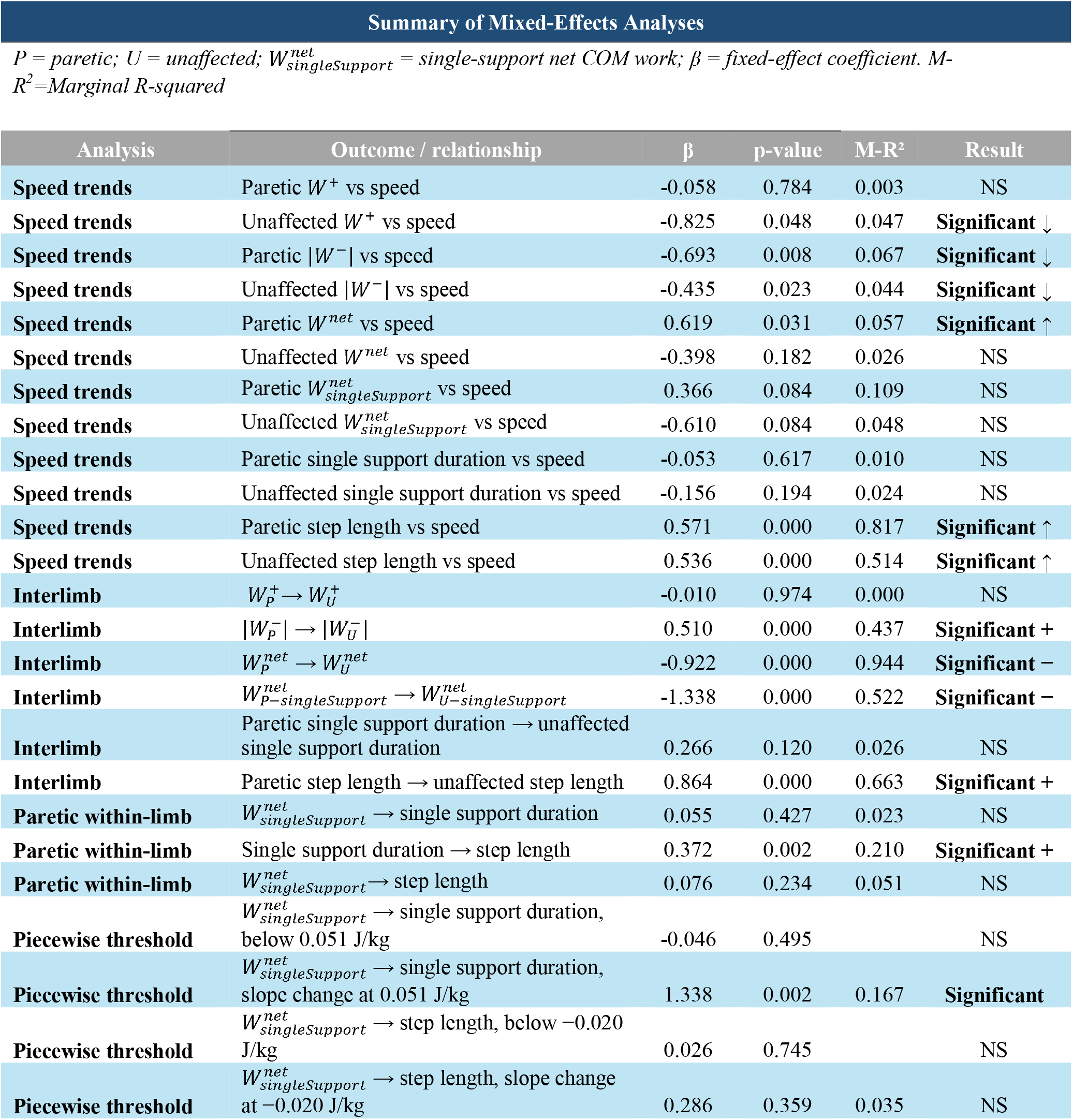
Summary of linear mixed-effects model (LMM) analyses of speed-dependent, interlimb, and within-paretic-limb relationships for COM mechanical work, single-support duration, and step length, including piecewise threshold analysis of paretic single-support net COM work. P, paretic limb; U, unaffected limb; *W* ^+^ positive COM work; | *W*^−^ | magnitude of negative COM work; *W* ^*net*^, net COM work; 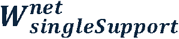, single-support net COM work; single-support duration; β, fixed-effect coefficient; R^2^, marginal coefficient of determination.

*Step length* increased significantly with walking speed for both limbs. Paretic step length increased from 0.18 ± 0.04 to 0.45 ± 0.03 m (β = 0.571, p < 0.001, marginal R^2^ = 0.817), while unaffected step length increased from 0.26 ± 0.08 to 0.47 ± 0.03 m (β = 0.536, p < 0.001, marginal R^2^ = 0.514).

*Positive COM work (W*^+^) showed no significant association with speed for the paretic limb (β = −0.058, p = 0.784, marginal R^2^ = 0.003), whereas unaffected *W*^+^ decreased significantly with speed (β = −0.825, p = 0.048, marginal R^2^ = 0.047), decreasing from 1.18 ± 0.69 to 0.36 ± 0.12 J·kg□^1^.

*Negative COM work (*|*W*^−^|) decreased significantly with speed for both limbs. Paretic |w^−^| decreased from 0.89 ± 0.56 to 0.28 ± 0.09 J·kg□^1^ (β = −0.693, p = 0.008, marginal R^2^ = 0.067), while unaffected | *W*^−^| decreased from 0.51 ± 0.24 to 0.13 ± 0.03 J·kg□^1^ (β = −0.435, p = 0.023, marginal R^2^ = 0.044).

*Net COM work (W*^*net*^) increased significantly with speed for the paretic limb, from −0.63 ± 0.61 to −0.06 ± 0.16 J·kg□^1^ (β = 0.619, p = 0.031, marginal R^2^ = 0.057). Unaffected *W*^*net*^ showed a negative but nonsignificant association with speed (β = −0.398, p = 0.182, marginal R^2^ = 0.026), decreasing descriptively from 0.67 ± 0.56 to 0.24 ± 0.11 J·kg□^1^.

*Single-support net COM work* 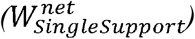 showed no significant linear association with speed for either limb. The paretic limb showed a positive association that did not reach significance (β = 0.366, p = 0.084, marginal R^2^ = 0.109), whereas the unaffected limb showed a negative association (β = −0.610, p = 0.084, marginal R^2^ = 0.048).

*Single-support duration* showed no significant association with speed for either the paretic (β = −0.053, p = 0.617, marginal R^2^ = 0.010) or unaffected limb (β = −0.156, p = 0.194, marginal R^2^ = 0.024).

### Interlimb relationships

Interlimb LMM analysis showed no relationship between paretic and unaffected *W*^+^ (β = −0.010, p = 0.974, marginal R^2^ < 0.001). In contrast, | *W*^−^| was positively associated between limbs (β = 0.510, p < 0.001, marginal R^2^ = 0.437). *W*^*net*^ showed a strong inverse interlimb relationship (β = −0.922, p < 0.001, marginal R^2^ = 0.944), and 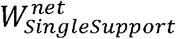 was also inversely related between limbs (β = −1.338, p < 0.001, marginal R^2^ = 0.522).

Single support duration was not significantly associated between limbs (β = 0.266, p = 0.120, marginal R^2^ = 0.026), whereas step length showed a strong positive interlimb relationship (β = 0.864, p < 0.001, marginal R^2^ = 0.663). Within the paretic limb, single support duration was positively associated with step length (β = 0.372, p = 0.002, marginal R^2^ = 0.210). In contrast, 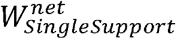 was not linearly associated with either single support duration (β = 0.055, p = 0.427, marginal R^2^ = 0.023) or step length (β = 0.076, p = 0.234, marginal R^2^ = 0.051).

### Single-support net-work threshold

Piecewise LMM analysis identified a breakpoint at 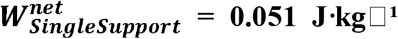 for the relationship between paretic 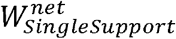 and single support duration. Below this breakpoint, the relationship was not significant (β = −0.046, p = 0.495). Above the breakpoint, the slope increased to 1.292, with a significant change in slope (Δβ = 1.338, p = 0.0018; marginal R^2^ = 0.167).

For step length, the piecewise model identified a breakpoint at −0.020 J·kg□^1^, but neither the relationship below the breakpoint (β = 0.026, p = 0.745) nor the change in slope (Δβ = 0.286, p = 0.359; marginal R^2^ = 0.035) was significant. Thus, a significant threshold relationship was observed specifically between paretic 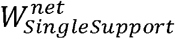 and single support duration.

## DISCUSSION

Human walking speed is not determined solely by propulsion magnitude but by how the neuromechanical system organizes mechanical energy across stance, transition, and single-support phases. In healthy walking, active work is temporally concentrated: trailing-limb push-off is timed to reduce leading-limb collision losses during the step-to-step transition^5,7^. The required push-off work scales geometrically with walking speed and step length^5^; when push-off work magnitude and timing satisfy the redirection demand^9^, collision is minimized and net single-support work remains near zero^6,18^.

In this study, the simulation provides a mechanical reference for determining how insufficient push-off reorganizes work across the step. Rather than predicting experimental responses from walking speed, the model identifies the consequences of progressive push-off deficiency, including redistribution of positive work before or after the step transition, step-length adjustment, and alteration of the COM trajectory. Each walking-speed condition was therefore evaluated independently as a distinct mechanical state, with COM power form used to identify the corresponding push-off deficiency regime. Empirical work patterns were then compared with the compensatory strategies predicted by the simulation. Accordingly, the simulation speed was not used to scale or directly predict experimental outcomes; the simulation was used to establish mechanistic predictions that were evaluated independently at each empirical walking speed. Accordingly, below we distinguish between model predictions, empirical observations, and whether the observed patterns are consistent with those predictions; the empirical observations are not interpreted as direct validation of the model but are related to the factors exposed by the model.

The observed hemiparetic COM power patterns differ from the phase-concentrated organization previously reported in able-bodied walking (**FIGURE 6**). The COM power form indicates that when paretic push-off is insufficient ^17^, redirection work cannot be confined to the transition window and active work redistributes across the step including single stance ^6,9,18^.

In healthy walking, early stance exhibits a distinct negative power peak reflecting a collision-like redirection of the COM velocity by the leading limb heel-strike ^7,17^. At the lowest walking speeds, however, the paretic limb frequently lacks clear early-stance collision-related negative power. This attenuation must be associated with limited load acceptance rather than with absence of heel strike ^33,34^: if weight support capacity is insufficient, the limb does not generate a substantial collision impulse and therefore it does not express collision mechanics in the same way as the unaffected limb^23^. Under such conditions, the absence of paretic-side COM redirection is consistent with the model-predicted extreme push-off condition, in which COM redirection is completed prior to paretic stance Hence, it indicates that the nonparetic push-off work fully eliminates the redirection demand from the paretic leg by shifting the temporal burden of redirection to late stance of the unaffected leg. Consequently, the paretic leading limb encounters a COM trajectory that has already been redirected, eliminating the need for the collision impulse ^7,9^ and altering the overall vertical COM oscillation pattern (**FIGURE 1**D and **FIGURE 7**). Therefore, the collision-eliminating push-off impulse must be twice as large as the associated optimal impulse and as a result elevates the step transition work to four times the optimal magnitude^7^. The small early-stance negative-power fluctuations observed on the paretic side are therefore more plausibly attributed to contact transients and event detection attempts rather than to healthy, optimized collision. In post-stroke gait, altered ankle stiffness and heightened reflex gains may amplify these early-stance transients, potentially triggering rapid initiation of positive work^35^.

When late-stance paretic push-off work is extremely diminished or absent, early-stance positive work is observed before the step transition, consistent with the pre-transition work redistribution predicted by the model, redistributing redirection work into adjacent phases rather than eliminating it. Conversely, when limited paretic push-off work is present, a prominent rebound phase on the unaffected side appears after transition, reflecting *post-transition compensation*^9,18^. Thus, depending on residual paretic capacity, positive work may be redistributed before the transition, whereas increased rebound work may provide post-transition compensation, but in both cases the defining feature is temporal redistribution of step positive work (*W*^+^) across the step.

Consistent with the extreme pre-emptive push-off regime, the vertical COM trajectory demonstrates a structural reorganization of step dynamics when paretic push-off work is absent. In this condition, unaffected limb heel-strike occurs at the vertical COM minimum, after which the COM rises throughout the unaffected limb stance and passes its maximum at paretic limb heel-strike. Immediately following paretic limb heel-strike, the COM begins descending without exhibiting a distinct collision-related redirection^7,17^. This sequence is consistent with the model-predicted extreme push-off condition, in which COM redirection is completed prior to paretic-limb contact^9^, effectively satisfying the geometric redirection requirement before the typical transition window^7^.

As a consequence, the paretic limb does not experience the ascending half of a pendular arc; instead, it inherits the COM at its vertex and undergoes only the descending phase of motion. At the population-average level, this produces one apparent rise–fall COM excursion per stride, rather than the two excursions expected from the two pendular steps comprising a stride ^36^. Such a reduction cannot arise from reduced propulsion magnitude alone; it reflects a collapse of bilateral pendular symmetry^36^ and the emergence of a single-limb–dominated oscillator in which the COM rises predominantly during unaffected-limb stance and descends during paretic-limb stance, a fundamental change in the dynamic strategy of providing support and progress.

The absence of a collision-based negative power phase, therefore, does not imply elimination of redirection demand, but rather its temporal relocation to late stance of the unaffected limb. Importantly, operating near this collision-eliminating regime implies that hemiparetic walking without paretic push-off work is mechanically extreme: the nonparetic limb must generate substantial positive work, even at low speeds, rendering the gait energetically demanding despite its reduced velocity. The vertical trajectory evidence, therefore, also provides geometric confirmation that, under severe paretic push-off work deficiency, active mechanical work is no longer transition-concentrated but is performed in another phase of the step, fundamentally altering whole-body energetic organization The vertical COM trajectory therefore provides additional evidence of altered COM motion when paretic push-off work is severely diminished or absent, with the observed trajectory being consistent with redistribution of positive mechanical work outside the typical transition phase^19^.

Based on our simulation, we anticipated a more flattened COM trajectory (approaching horizontal translation) over paretic limb stance as a consequence of altered redirection mechanics. However, this trajectory was observed specifically in the condition where paretic push-off work was absent and the paretic limb exhibited substantial negative work during unaffected limb stance. This indicates that the altered COM vertical profile is unlikely to reflect an intentional control strategy aimed at modulating step transition demand for the paretic limb side. Rather, it is more parsimoniously interpreted as being consistent with the mechanical consequences of net energy dissipation during paretic stance that reduces COM kinetic energy, limiting vertical excursion and effectively flattening the COM path during the subsequent unaffected limb stance.

Net step work results reveal a persistent energetic asymmetry. Paretic limb net work remained negative across speeds (−0.63 −0.06 → J.kg^− 1^), while unaffected limb net work remained positive (0.67 → 0.24 J.kg^−1^). This opposing pattern of limb-specific net COM work reflects a redistribution of mechanical work between the limbs. From an optimization viewpoint, two individuals with similar paretic propulsion may differ fundamentally in timing organization, stance dissipation, and the compensation phase. This suggests that mechanical work organization may reflect constrained optimization, with work distributed across limbs and gait phases according to the constraints imposed on the locomotor system ^19^. Thus, propulsion alone cannot characterize energetic organization in the gait.

The inverse relationship in limb-specific net COM work was substantially stronger than the relationships between limb-specific net COM work and walking speed. Whole-step net work demonstrated an exceptionally strong inverse interlimb association (β = −0.922, marginal R^2^ = 0.944), whereas speed explained comparatively little variance in paretic and unaffected net work (marginal R^2^ = 0.057 and 0.026, respectively). A similar pattern emerged during single support: net work showed a substantial inverse interlimb association (β = −1.338, marginal R^2^ = 0.522), despite neither limb exhibiting a significant linear association with speed. Positive work showed essentially no interlimb relationship, whereas negative work was positively associated between limbs, indicating that the strongest reciprocal organization emerged specifically in the **net energetic balance between the limbs**. This reciprocity is mechanically consequential because sustained walking requires whole-body mechanical energy balance: persistent net energy removal by one limb must ultimately be offset by net energy addition elsewhere. Thus, walking speed describes the locomotor performance achieved, whereas the reciprocal interlimb work relationship more directly characterizes **how the energetic demands of that performance are organized between the limbs**.

Step length increased strongly with speed for both limbs, yet paretic step length remained shorter at matched speeds. From the perspective of step-transition mechanics, the required trailing-limb push-off work increases with step length^5,7,17^; reducing paretic step length therefore reduces the subsequent push-off work demand^7^. However, the present results indicate that step length should not be interpreted solely as an adaptation to match reduced step-transition work capacity. Step length is an integrated outcome of both the transition work demand and the mechanical conditions under which the preceding single-support phase can be sustained. Consequently, step-length asymmetry alone cannot distinguish whether a shorter parameter reflects reduced transition work capacity, a constraint arising during single support, or a combination of both.

The simulation provides a second potential pathway through which step length may be constrained, arising during single support rather than the step transition (**FIGURE 5**). Net removal of mechanical energy during single support progressively reduces COM kinetic energy. If sufficient energy is removed before completion of the intended pendular progression, COM velocity approaches zero and continuation of the step becomes mechanically infeasible. In the simulation, this condition necessitates earlier contralateral heel strike and consequently reduces achievable step length.

The empirical results reveal an important departure from the single-support behavior predicted by the pendular simulation. Paretic single-support duration was significantly associated with paretic step length (β = 0.372, p = 0.002, marginal R^2^ = 0.210), whereas single-support net work was not linearly associated with either duration or step length. Piecewise analysis, however, identified a transition near 0.05 J·kg□^1^, close to the energy-neutral condition. Above this boundary, greater single-support net work was associated with longer single-support duration; below it, the relationship disappeared. This differs from the perturbed pendulum model, in which progressively greater opposing work continues to prolong single-support motion until a critical energetic threshold is reached, after which progression collapses. Thus, once paretic single support approaches an energy-neutral or net-dissipative state, its duration no longer follows the work-dependent progression predicted by the pendular model. The significant association between single-support duration and step length, despite the absence of a direct single-support work–step-length relationship, indicates that the temporal termination of paretic single support becomes an independent constraint on achievable step length.

Some specific limitations should be considered in this study. The first limitation of this study is the relatively small sample size (N = 11), which limits the generalizability of the findings. Although the repeated-measures design enabled evaluation across multiple walking speeds, the proposed mechanistic framework should be validated in larger and more diverse post-stroke cohorts. Second, all energetic quantities were derived from GRFs rather than joint-level inverse dynamics ^1^ or muscle activity level (EMG) ^37^. COM work reflects dominant whole-body mechanical action ^27^ but cannot uniquely attribute work to specific muscles or joints. Negative COM work does not necessarily imply passive collapse; internal muscle activity may coexist with external energy dissipation ^38^. Third, work partitioning depends ^8,9^ on event detection and phase boundary definitions. Although the redistribution pattern is robust, exact allocation across sub-phases may vary slightly with detection thresholds. Fourth, treadmill walking enabled controlled speed sweeps but may differ subtly from overground gait ^39^. Therefore, replication in overground contexts is warranted. And finally, causal inference cannot be established from cross-sectional speed comparisons alone. Experimental manipulation of paretic limb push-off work will be required to confirm whether restoring these capacities re-concentrates active work within healthy transition timing.

We also found a complementary limitation. The adapted analytical model quantifies the step-transition work required for optimal, suboptimal, or absent push-off at a prescribed walking speed. It also predicts that optimal step-transition work increases quadratically with walking speed. However, the empirical data showed that measured limb-specific positive COM work declined as participants walked faster. The model therefore cannot explain this observed speed-dependent reduction in mechanical work. Accordingly, the model should be interpreted as describing the mechanical consequences of push-off work limitation at a given speed, rather than predicting the full speed dependence of hemiparetic COM work.

The present findings challenge the interpretation of walking speed as a stand-alone metric of locomotor recovery. Although walking speed quantifies the functional performance achieved, it provides limited information regarding the mechanical organization through which that performance is produced. This distinction was particularly evident for net COM work: its association with walking speed was weak for both the paretic and unaffected limbs, whereas paretic and unaffected net work exhibited an exceptionally strong reciprocal relationship. Thus, individuals walking at a given speed may achieve that performance through fundamentally different distributions of mechanical responsibility between the limbs. An increase in walking speed therefore cannot, by itself, distinguish restoration of paretic-limb mechanical capacity from greater mechanical work contribution by the unaffected limb. The phase- and limb-specific measures presented here provide complementary mechanistic outcomes by identifying whether improved walking performance results from restoration of paretic contribution, redistribution of work to the unaffected limb, altered single-support capacity, or combinations of these adaptations. Accordingly, walking speed may describe the level of locomotor performance, whereas the present framework evaluates the mechanical basis of recovery.

Hemiparetic walking across the examined speeds is characterized not merely by reduced propulsion, but by a fundamental redistribution of active mechanical work across the entire step ^9^. When paretic trailing-limb push-off is insufficient relative to the redirection demand, transition work cannot remain specific phase-concentrated ^7^ and instead disperses into early stance and single support ^1,9^. Elevated paretic negative work and persistently negative paretic net COM work indicate a mechanical work distribution in which the paretic limb shows net energy removal, whereas the nonparetic limb shows net energy addition.

The single-support simulation identifies a mechanical threshold at which progressive energy removal can terminate pendular progression; however, the empirical data show that paretic single-support behavior does not follow this work-dependent prediction once net work approaches the energy-neutral or dissipative regime. Instead, single-support duration is directly associated with step length, whereas single-support net work is not. The significant piecewise work–duration transition therefore identifies a departure from the work-dependent behavior predicted by the pendular model rather than a direct energetic determination of step length.

By integrating step transition mechanics^7^ with phase-resolved COM work partitioning ^9^, this framework extends classical step-to-step theory into pathological gait and advances beyond propulsion-only metrics. These findings motivate the hypothesis that future rehabilitation strategies targeting paretic-limb propulsion may also benefit from addressing stance-phase energy dissipation and its relationship to the temporal distribution of active work.

## Data Availability

All data produced in the present study are available upon reasonable request to the authors

## ACKNOLEDGEMENTS

We would like to thank Alexa Boyer, Dr. Deepthi Rajasekhar, Dr. Kelly Kaiser, Dr. Sean Dukelow, Dr. Gregor Kuntze, and the Physiotherapists of Unit 58 in the Foothills Hospital.

## DECLARATIONS

### Consent to participate

All participants signed an informed consent prior to their participation in this research.

### Ethics approval

This research is approved by ethics certificate #REB21-1576 issued by the University of Calgary Conjoint Health Research Ethics Board.

### Data availability

Data are available at https://github.com/salehhosseini/Hemiparetic-dataset

### Author contributions

Concept, planning, analysis, writing and editing - Seyed-Saleh Hosseini-Yazdi; Concept, planning, editing – John EA Bertram; Data collection, protocol design, editing – Karson Fitzsimons.

## REFERENCES

1. Farris, D. J., Hampton, A., Lewek, M. D. & Sawicki, G. S. Revisiting the mechanics and energetics of walking in individuals with chronic hemiparesis following stroke: from individual limbs to lower limb joints. J NeuroEngineering Rehabil 12, 24 (2015).

2. Alexander, R. M. Simple models of walking and jumping. Human Movement Science 11, 3–9 (1992).

3. Alexander, R. McN. Simple Models of Human Movement. Applied Mechanics Reviews 48, 461–470 (1995).

4. Cavagna, G. A., Heglund, N. C. & Taylor, C. R. Mechanical work in terrestrial locomotion: two basic mechanisms for minimizing energy expenditure. Am J Physiol 233, R243–261 (1977).

5. Donelan, J. M., Kram, R. & Kuo, A. D. Mechanical work for step-to-step transitions is a major determinant of the metabolic cost of human walking. J Exp Biol 205, 3717–3727 (2002).

6. Hosseini-Yazdi, S.-S. & Bertram, J. E. A. Center of mass work analysis predicts preferred walking speeds for varying walking conditions. Journal of Biomechanics 185, 112682 (2025).

7. Kuo, A. D. Energetics of actively powered locomotion using the simplest walking model. J Biomech Eng 124, 113–120 (2002).

8. Kuo, A. D., Donelan, J. M. & Ruina, A. Energetic Consequences of Walking Like an Inverted Pendulum: Step-to-Step Transitions: Exercise and Sport Sciences Reviews 33, 88–97 (2005).

9. Hosseini-Yazdi, S.-S. & Bertram, J. E. A. Optimum Push-off for Uneven Walking Based on the Just-in-Time Strategy: Walking with Interrupted Push-Off is Mechanically Costly. Journal of Biomechanical Engineering 1–20 (2025) doi:10.1115/1.4069666.

10. Hosseini-Yazdi Seyed-Saleh. Energetics and Biomechanics of Uneven Walking for Young and Older Adults. (University of Calgary, Calgary, AB, Canada, 2024).

11. Huang, T. P., Shorter, K. A., Adamczyk, P. G. & Kuo, A. D. Mechanical and energetic consequences of reduced ankle plantarflexion in human walking. Journal of Experimental Biology https://doi.org/10.1242/jeb.113910 (2015) doi:10.1242/jeb.113910.

12. Zelik, K. E. & Kuo, A. D. Human walking isn’t all hard work: evidence of soft tissue contributions to energy dissipation and return. Journal of Experimental Biology 213, 4257–4264 (2010).

13. Perry, J., Garrett, M., Gronley, J. K. & Mulroy, S. J. Classification of Walking Handicap in the Stroke Population. Stroke 26, 982–989 (1995).

14. Peterson, C. L., Hall, A. L., Kautz, S. A. & Neptune, R. R. Pre-swing deficits in forward propulsion, swing initiation and power generation by individual muscles during hemiparetic walking. J Biomech 43, 2348–2355 (2010).

15. Bowden, M. G., Balasubramanian, C. K., Neptune, R. R. & Kautz, S. A. Anterior-Posterior Ground Reaction Forces as a Measure of Paretic Leg Contribution in Hemiparetic Walking. Stroke 37, 872–876 (2006).

16. Patterson, K. K., Gage, W. H., Brooks, D., Black, S. E. & McIlroy, W. E. Evaluation of gait symmetry after stroke: A comparison of current methods and recommendations for standardization. Gait & Posture 31, 241–246 (2010).

17. Adamczyk, P. G. & Kuo, A. D. Redirection of center-of-mass velocity during the step-to-step transition of human walking. J Exp Biol 212, 2668–2678 (2009).

18. Hosseini-Yazdi, S.-S. & Bertram, J. E. The consequence of uneven walking transitory modulation strategies: A simulation-based approach. J Theor Biol 614, 112234 (2025).

19. Bertram, J. E. A. Constrained optimization in human walking: cost minimization and gait plasticity. Journal of Experimental Biology 208, 979–991 (2005).

20. Kuo, A. D. A simple model of bipedal walking predicts the preferred speed-step length relationship. J Biomech Eng 123, 264–269 (2001).

21. Darici, O., Temeltas, H. & Kuo, A. D. Optimal regulation of bipedal walking speed despite an unexpected bump in the road. PLoS ONE 13, e0204205 (2018).

22. Donelan, J. M., Kram, R. & Kuo, A. D. Simultaneous positive and negative external mechanical work in human walking. J Biomech 35, 117–124 (2002).

23. Ruina, A., Bertram, J. E. A. & Srinivasan, M. A collisional model of the energetic cost of support work qualitatively explains leg sequencing in walking and galloping, pseudo-elastic leg behavior in running and the walk-to-run transition. Journal of Theoretical Biology 237, 170–192 (2005).

24. Darici, O. & Kuo, A. D. Humans optimally anticipate and compensate for an uneven step during walking. eLife 11, e65402 (2022).

25. Darici, O., Temeltas, H. & Kuo, A. D. Anticipatory Control of Momentum for Bipedal Walking on Uneven Terrain. Sci Rep 10, 540 (2020).

26. Hosseini-Yazdi, S.-S. & Kuo, A. D. The energetic cost of human walking as a function of uneven terrain amplitude. Journal of Experimental Biology jeb.249840 (2025) doi:10.1242/jeb.249840.

27. Van Der Zee, T. J. & Kuo, A. D. Soft tissue deformations explain most of the mechanical work variations of human walking. Journal of Experimental Biology 224, jeb239889 (2021).

28. Bertram, J. E. A., D’Antonio, P., Pardo, J. & Lee, D. V. Pace length effects in human walking: ‘groucho’ gaits revisited. J Mot Behav 34, 309–318 (2002).

29. Kajita, S. & Tani, K. Study of dynamic biped locomotion on rugged terrain-derivation and application of the linear inverted pendulum mode. in Proceedings. 1991 IEEE International Conference on Robotics and Automation 1405–1411 (IEEE Comput. Soc. Press, Sacramento, CA, USA, 1991). doi:10.1109/ROBOT.1991.131811.

30. Müller, R. & Blickhan, R. Running on uneven ground: leg adjustments to altered ground level. Hum Mov Sci 29, 578–589 (2010).

31. Müller, R., Ernst, M. & Blickhan, R. Leg adjustments during running across visible and camouflaged incidental changes in ground level. Journal of Experimental Biology 215, 3072–3079 (2012).

32. Day, E. M., Alcantara, R. S., McGeehan, M. A., Grabowski, A. M. & Hahn, M. E. Low-pass filter cutoff frequency affects sacral-mounted inertial measurement unit estimations of peak vertical ground reaction force and contact time during treadmill running. J Biomech 119, 110323 (2021).

33. Chen, G., Patten, C., Kothari, D. H. & Zajac, F. E. Gait differences between individuals with post-stroke hemiparesis and non-disabled controls at matched speeds. Gait & Posture 22, 51–56 (2005).

34. Olney, S. J. & Richards, C. Hemiparetic gait following stroke. Part I: Characteristics. Gait & Posture 4, 136–148 (1996).

35. Hsiao, H.-Y., Gray, V. L., Borrelli, J. & Rogers, M. W. Biomechanical control of paretic lower limb during imposed weight transfer in individuals post-stroke. J NeuroEngineering Rehabil 17, 140 (2020).

36. Winter, D. A. Biomechanics and Motor Control of Human Gait. (Univ. of Waterloo Press, Waterloo, Ontario, 1991).

37. Souissi, H., Zory, R., Bredin, J., Roche, N. & Gerus, P. Co-contraction around the knee and the ankle joints during post-stroke gait. Eur J Phys Rehabil Med 54, (2018).

38. Balbinot, G., Schuch, C. P., Bianchi Oliveira, H. & Peyré-Tartaruga, L. A. Mechanical and energetic determinants of impaired gait following stroke: segmental work and pendular energy transduction during treadmill walking. Biol Open 9, bio051581 (2020).

39. Riley, P. O., Paolini, G., Croce, U. D., Paylo, K. W. & Kerrigan, D. C. A kinematic and kinetic comparison of overground and treadmill walking in healthy subjects. Gait Posture 26, 17–24 (2007).

